# Longitudinal subcortical volume changes and their correlations with multiple PET and fluid biomarkers in dominantly inherited Alzheimer’s disease

**DOI:** 10.1101/2025.11.23.25340829

**Authors:** IL Han Choo, Hoyoung Park, the Dominantly Inherited Alzheimer Network

## Abstract

**Objective:** To investigate longitudinal subcortical structural changes in autosomal dominant Alzheimer’s disease in relation to multiple PET and fluid biomarkers.

**Methods:** Participants underwent structural MRI, ^11^C-Pittsburgh Compound B PET, ^18^F-fluorodeoxyglucose PET, and CSF and plasma assessments. Rates of biomarker change as a function of estimated years to symptom onset were estimated using multivariate linear mixed-effects models, and longitudinal associations between subcortical atrophy and multiple biomarkers were evaluated.

**Results:** A total of 601 participants completed one or more clinical evaluations, with up to eight annual visits. Mutation carriers showed significantly greater longitudinal atrophy in the left amygdala, bilateral thalamus, putamen, nucleus accumbens, and hippocampus compared with non-carriers (Bonferroni-corrected p < 0.05). The earliest divergence was observed 13.2 years before the expected symptom onset in the right nucleus accumbens, following amyloid-β (Aβ) accumulation in the right thalamus that began 23.8 years before onset. Among carriers, atrophy in the right thalamus, bilateral putamen, and bilateral nucleus accumbens was significantly associated with region-specific or cortical Aβ accumulation, as well as with CSF Aβ42, Aβ42/Aβ40 ratio, total tau, and phosphorylated tau (Bonferroni-corrected p < 0.05).

**Conclusion:** As MRI is more cost-effective and time-efficient than PET, subcortical volume atrophy measured by MRI may serve as a practical biomarker for the early detection of Alzheimer’s disease.

## INTRODUCTION

Alzheimer’s disease dementia (AD) is most common cause of dementia worldwide. Studies of individuals from families with autosomal dominant Alzheimer’s disease (ADAD) provide a unique opportunity to characterize the earliest pathophysiological changes that occur before symptom onset. While magnetic resonance imaging (MRI) investigations in AD have primarily focused on the cerebral cortex and reported only cross-sectional findings,^1,2^postmortem studies demonstrate that amyloid plaques and neurofibrillary tangles are also present in subcortical regions.^3,4^ Consistently, amyloid positron emission tomography (PET) studies in presymptomatic familial AD have identified the striatum and thalamus as early sites of amyloid deposition,^5^ and reductions in subcortical volume have been associated with both clinical severity and cognitive decline.^6^

Findings from the Dominantly Inherited Alzheimer Network (DIAN) further highlight the importance of these regions: significant gray matter volume differences between noncarriers and mildly symptomatic carriers (Clinical Dementia Rating [CDR] 0.5) have been observed in the thalamus and putamen.^7^ However, subsequent longitudinal DIAN studies have not focused on the difference in rate of subcortical volume change between carriers and non-carriers.^8,9^ This gap in knowledge is clinically relevant, as MRI—widely used in routine diagnostic practice—could provide a cost- and time-efficient means to detect such changes. If MRI-based measures of subcortical atrophy can be incorporated into early diagnostic strategies, analogous to plasma phospho-tau217,^10^ they could facilitate broader population screening than methods based on PET or cerebrospinal fluid (CSF) analysis.

To date, no study has comprehensively investigated longitudinal subcortical structural changes in ADAD in relation to multiple PET and fluid biomarkers. We therefore hypothesized that (1) the rates of a priori–selected subcortical regional atrophy differ longitudinally, relative to estimated years to symptom onset, between mutation carriers and non-carriers; and (2) subcortical regions showing significant longitudinal differences by mutation status also demonstrate longitudinal associations with amyloid burden (^11^C-Pittsburgh compound B [PIB] PET), glucose metabolism (^18^F-fluorodeoxyglucose [FDG] PET), and fluid biomarkers (CSF Aβ42, Aβ42/Aβ40, CSF Tau, CSF pTau, Plasma Aβ1-42, Plasma Aβ1-42/Aβ1-40).

## METHODS

### Study design and participants

Individuals were included from the DIAN Observational Study (the 16th semiannual data freeze). Participants were identified as mutation-carriers (MCs) of pathologic variants in presenilin-1 (PSEN1), presenilin-2 (PSEN2), or amyloid precursor protein (APP) and as non-carriers (NCs) from the same families as the MCs. They underwent baseline and 2 and more times longitudinal follow-up assessments of PIB/FDG-PET, MRI, CSF Aβ42, CSF Aβ40, CSF Tau, CSF pTau, Plasma Aβ1-42, Plasma Aβ1-40.

DIAN estimated years to symptom onset (EYO) was calculated for symptomatic individuals with reported decline ages, their EYO = visit age – the mean of decline ages and for asymptomatic individuals, their EYO = their mutation/parental EYO.

Cognitive status was determined using the Clinical Dementia Rating (CDR) scale.^11^ A CDR of 0 for NCs defines cognitive normal (NC_CN). MC participants with CDR = 0 were identified as “presymptomatic” (MC_p), while subjects with CDR > 0 were identified as “symptomatic” which were divided as NCs with mild cognitive impairment (NC_MCI), MCs with mild cognitive impairment (MC_MCI) and MCs with AD (MC_AD). Apolipoprotein E (APOE) ε4 status was dichotomized in all participants between ε4 carriers (E4+, including heterozygotes and homozygotes) and ε4 non-carriers (E4-) (Table 1).

**Table 1.** Demography and Clinical Information.

|  | NC_CN<br>(n = 221) | NC_MCI<br>(n = 16) | MC_p<br>(n = 236) | MC_MCI<br>(n = 64) | MC_AD<br>(n = 64) |
| --- | --- | --- | --- | --- | --- |
| Age, Mean (SD) | 37.49 (11.04) | 40.68 (13.03) | 34.20 (9.16) | 42.94 (10.12) | 48.81 (8.97) |
| Female % | 58.82 | 56.25 | 58.05 | 59.38 | 43.75 |
| Estimated years of<br>symptom onset, Mean<br>(SD) | 48.76 (6.33) | 48.41 (4.77) | 48.51 (6.81) | 45.54 (6.39) | 45.16 (7.97) |
| CDR, Mean (SD) | 0.00 (0.00) | 0.47 (0.12) | 0.01 (0.10) | 0.49 (0.11) | 1.13 (0.73) |
| Family mutation<br>PSEN1/PSEN2/APP (%) | 70/10/20 | 56/0/44 | 70/11/19 | 80/<5/19 | 84/2/14 |
| APOE E4+ (number, %) | 69 (31.22%) | 5 (31.25%) | 70 (29.56%) | 17 (26.56%) | 19 (29.69%) |
| APOE E4- (number, %) | 152 (68.78%) | 11 (68.75%) | 166 (70.34%) | 47 (73.44%) | 45 (70.31%) |
| Visit distribution | {1: 78, 2: 64, 3: 39, 4: 23,<br>5: 20, 6: 5} | {1: 13, 3: 4, 4: 1} | {1: 76, 2: 83, 3: 36, 4: 23,<br>5: 18, 6: 4, 8: 1} | {1: 32, 2: 26, 3: 8, 4: 6, 6: 1} | {1: 20, 2: 34, 3: 14, 4: 6,<br>6: 2} |
| Follow up Diagnostic<br>Change | NC → NC_MCI: 4<br>NC → NC_MCI → NC: <5 | NC_MCI → NC: 7<br>NC_MCI → NC → NC_MCI:<br><5 | MC_p → MC_MCI: 14<br>MC_p → MC_MCI → MC_p: 3<br>MC_p → MC_MCI → MC_AD: 5<br>MC_p → MC_AD: <5 | MC_MCI → MC_AD: 21<br>MC_MCI → MC_p: 6<br>MC_MCI → MC_p → MC_MCI: <5<br>MC_MCI → MC_p → MC_MCI →<br>MC_AD: <5<br>MC_MCI → MC_p → MC_AD: <5 | MC_AD → MC_MCI: <5 |

Each Diagnostic group and number were categorized by baseline visit.
Visit distribution was displayed by visit count: distinct numbers of ID. These distinct ID numbers were also used in follow up diagnostic change counts.

DIAN participants provided informed consent in accordance with the local institutional review boards of each participating site. DIAN study procedures have received ethics approval by the Human Research Protection Office at Washington University in St. Louis (MO, USA) and all of the participating sites. Additional institutional review board approval (CHOSUN 2024-08-019-002) was received from Chosun University Hospital for data analysis.

#### MRI Imaging and Processing

Structural MRI acquisition was performed using the Alzheimer Disease Neuroimaging Initiative (ADNI) protocol.^12,13^ Participating sites were required to pass initial and regular follow-up quality control assessments to insure acquisition conformity. Each participant received an accelerated 3D sagittal T1-weighted MPRAGE on a 3T scanner. A high quality, whole-brain image with 1.1×1.1×1.2 mm voxels was acquired in approximately 5-6 minutes. Before analysis, images were screened for artifacts and protocol compliance by the ADNI imaging core. FreeSurfer analysis involves subcortical reconstruction and volumetric segmentation of T1 weighted images.^14^ MRI regional volumes were corrected for head size (intracranial volume, ICV) in order to have correct comparisons as follows: First, Compute mean ICV for sample; Second, Compute regression with ICV as independent variable and an ROI as dependent variable to obtain B weight; Third, Compute: Normalized = raw volume – (B-weight * (ss ICV – mean ICV)) (“ss” = single subject’s).

#### PET Processing

Each site underwent an initial evaluation by the ADNI PET QC site to ensure compliance with a common PIB, FDG PET protocol. Amyloid imaging was performed with a bolus injection of approximately 15 mCi of PIB. Dynamic imaging acquisition started either at injection for 70 minutes or 40 minutes post-injection for 30 minutes. For analysis, the PIB PET data between 40 to 70 minutes was converted to regional standardized uptake value ratios (SUVRs). Metabolic imaging with FDG-PET was performed with a 3D dynamic acquisition began 40 minutes after a bolus injection of approximately 5 mCi of FDG. Data from the last 20 minute of each FDG scan were converted to SUVRs relative to cerebellar grey matter. PET imaging analyses are performed using the PET unified pipeline.^15,16^ PET images are smoothed to achieve a common spatial resolution of 8mm to minimize inter-scanner differences.^17^ Inter-frame motion correction for the dynamic PET images is performed using standard image registration techniques.^18,19^ PET-MR registration is performed using a vector-gradient algorithm (VGM)^20^ in a symmetric fashion (i.e. average transformation for PET->MR and inverse of MR->PET was used as the final transformation matrix). By default, regional PET processing is performed based on FreeSurfer segmentation (using wmparc.mgz as the region definition), and each FreeSurfer region is analyzed. To assess global amyloid burden based on amyloid PET imaging data, the arithmetic mean SUVR from precuneus, prefrontal cortex, gyrus rectus, and lateral temporal regions are defined as the mean cortical SUVR.

#### Biofluids Analysis

Protocols for the collection of cerebrospinal fluid (CSF) and blood (for plasma) are consistent with the biofluid protocol of the Alzheimer’s Disease Neuroimaging Initiative (ADNI) (http://www.adni-info.org/). CSF amyloid β42 (Aβ42), tau (Tau), and phosphorylated tau-181 (pTau) were measured by immunoassay using Luminex bead-based multi-plexed xMAP technology (INNO-BIA AlzBio3, Innogenetics). Concentrations of CSF Aβ40 were measured by plate-based enzyme-link immunosorbant assay (ELISA) (research prototype INNOTEST™ Aβ40, Innogenetics, Ghent, Belgium). Aβ42 estimates were normalized for individual differences in CSF production rates by forming a ratio with Aβ40 as the denominator.^21^ Blood samples were collected via venipuncture under fasting conditions. Concentrations of plasma Aβ species (Aβ1-40, Aβ1-42) were measured by xMAP (INNO-BIA Plasma Aβ Forms Multiplex Assay™, Innogenetics, Ghent, Belgium).^22^

### Statistical analysis

All statistical analyses were performed using R version 4.5.0 (The R Foundation for Statistical Computing, http://www.r-project.org/). A two-step analytical framework was employed to test our hypotheses.

In the first step, we conducted reference analyses using multivariate linear mixed-effects models (LMMs) implemented via the lme4 (version 1.1-35.3) and lmerTest (version 3.1-3) packages. The models included fixed effects for estimated years from symptom onset (EYO), mutation status (autosomal dominant Alzheimer’s disease (ADAD) mutation carriers vs. non-carriers), and time from baseline, along with all possible two-way and three-way interaction terms. Sex, years of education, and APOE ε4 carrier status were considered as additional covariates. Each covariate was individually added to the baseline model—which already included all fixed effects and interaction terms—and evaluated using a likelihood ratio test via ANOVA. Only covariates that showed a statistically significant improvement in model fit (p < 0.05) were retained in the final model specification. The random effects included in the models were the random intercepts for family clusters, individual random intercept and random slope with unstructured covariance matrix, to account for the within-subject correlation due to repeated measures. These models were applied to examine the differential longitudinal trajectories of established AD biomarkers, including PIB and FDG PET measures, CSF Aβ42, CSF Aβ42/Aβ40 ratio, CSF Tau, CSF pTau, plasma Aβ1-42, and plasma Aβ1-42/Aβ1-40 ratio. In parallel, we examined longitudinal subcortical volume changes using the same LMM specification. Seven subcortical regions of interest (ROIs) — the amygdala, putamen, thalamus, caudate, pallidum, nucleus accumbens, and hippocampus — were each analyzed bilaterally, yielding a total of 14 ROI assessments based on a priori laterality hypotheses. Statistical significance was determined based on Bonferroni-adjusted p-values with a significance threshold of 0.05. For all significant findings, we further identified the specific EYO points at which significant group-level differences between mutation carriers and non-carriers began to emerge. Our primary statistical inference was based on the linear mixed-effects model estimates. For visualization purposes, we applied locally estimated scatterplot smoothing (LOESS) — a nonparametric technique effective for capturing nonlinear patterns — separately for each group to illustrate potential trends in the data. We interpreted the earliest time point at which the 99% confidence intervals of the LOESS-predicted trajectories for mutation carriers and non-carriers ceased to overlap as the onset of a statistically meaningful divergence.

In the second step, we explored whether the subcortical regions that showed significant longitudinal differences by mutation status in the first-step analysis were also longitudinally associated with the aforementioned AD biomarkers. Specifically, we assessed whether subcortical volume trajectories were linked to markers of amyloid burden (e.g., PIB SUVR), glucose metabolism (FDG SUVR), CSF Aβ42 and Aβ42/Aβ40, tau pathology (CSF Tau and pTau), and plasma Aβ species. For these LMM, we calculated the conditional coefficient of determination (R^2^c), which quantifies the variance explained by both fixed and random effects, using the MuMIn package (version 1.47.5). These analyses allowed us to evaluate the extent to which subcortical atrophy was coupled with multiple domains of AD pathology, particularly in mutation carriers.

## RESULTS

Demographic and clinical information on the participants is presented in Table 1. A total of 601 participants underwent one or more clinical evaluations with a maximum of eight visits and visit distribution details are provided in Table 1. Among them, 221 individuals were classified non-carriers with cognitive normal (NC_CN), and 236 individuals were presymptomatic mutation carriers (MC_p, CDR = 0) and 144 individuals were symptomatic, consisting of 16 non-carriers with mild cognitive impairment (NC_MCI), 64 mutation carriers with mild cognitive impairment (MC_MCI), and 64 mutation carriers with AD (MC_AD) by a CDR score of 0.5 or higher. For the analyses: MRI subcortical volume data were available for 566 individuals, comprising a total of 1330 observations. ^11^C-PIB PET and ^18^F-FDG PET data were available for 532 individuals, totaling 1130 observations. CSF Aβ42 and CSF Aβ42/Aβ40 ratio data were available for 462 individuals, totaling 844 observations. CSF Tau and CSF pTau data were available for 460 individuals, totaling 832 observations. Plasma Aβ1-42 and plasma Aβ1-42/Aβ1-40 ratio data were available for 517 individuals, comprising 1083 observations. Participants with longitudinal data had a mean 2.5 visits (SD 1.5) and 3.1 years (SD 3.0) of data.

### Longitudinal Association of AD biomarkers with Respect to EYO Between Mutation Carriers and Non-Carriers

Reference AD biomarker analyses revealed that mutation carriers exhibited significantly higher rates of increase compared to non-carriers in mean cortical PIB SUVR, CSF Tau, and CSF pTau, and significantly greater rates of decrease in precuneus FDG SUVR, CSF Aβ42, and CSF Aβ42/Aβ40 ratio. The estimated points of divergence between mutation carriers and non-carriers occurred at 22.5, 19.2, and 21.5 years before expected symptom onset for mean cortical PIB SUVR, CSF Tau, and CSF pTau, respectively, and at 12.0, 13.8, and 17.6 years before expected onset for precuneus FDG SUVR, CSF Aβ42, and CSF Aβ42/Aβ40 (Table 2, Figure 1). For subcortical regional volumes, 9 out of 14 subcortical ROIs — including the left amygdala, bilateral thalamus, putamen, nucleus accumbens, and hippocampus — demonstrated significant differences between mutation carriers and non-carriers. The earliest divergence was observed in the right nucleus accumbens at 13.2 years before expected symptom onset, with divergence across regions spanning from 13.2 years before to 4.5 years after expected onset (Table 3, Figure 2). Regarding ^11^C-PIB PET measures in subcortical regions, 11 out of 14 regions — including the right amygdala, bilateral thalamus, putamen, pallidum, caudate, and nucleus accumbens — showed significantly greater longitudinal rates of amyloid accumulation in mutation carriers relative to non-carriers. The estimated divergence points varied by region, ranging from 23.8 years to 15.7 years before expected symptom onset (Table 4, Figure 2). In contrast, for 18F-FDG PET measures, only the left hippocampus showed a significant decline in FDG SUVR between mutation carriers and non-carriers. Divergence occurred 8.1 years before the expected symptom onset in the left hippocampus (Table 4, Figure 2).

**Table 2.** Longitudinal linear mixed-effects model of reference AD biomarkers by estimated years to symptom onset interaction with mutation status.

|  | Fixed-effects<br>coefficient | SE | df | t-value | p-value |
| --- | --- | --- | --- | --- | --- |
| Mean Cortical PIB SUVR | 0.022 | 0.003 | 280.00 | 8.19 | **<0.001\*** |
| Precuneus FDG SUVR | -0.008 | 0.001 | 305.00 | -6.47 | **<0.001\*** |
| CSF Aβ42 | -21.087 | 3.154 | 252.00 | -6.69 | **<0.001\*** |
| CSF Aβ42/Aβ40 | -0.001 | 0.000 | 250.00 | -2.76 | **0.006\*** |
| CSF Tau | 3.526 | 0.585 | 253.00 | 6.03 | **<0.001\*** |
| CSF pTau | 1.955 | 0.262 | 252.00 | 7.45 | **<0.001\*** |
| Plasma Aβ1-42 | -0.023 | 0.142 | 540.00 | -0.16 | 0.871 |
| Plasma Aβ1-42/Aβ1-40 | 0.002 | 0.001 | 379.00 | 1.89 | 0.060 |

Mutation status are divided by mutation carriers and non-carriers of autosomal dominant Alzheimer's disease. \*p < 0.05; SE = Standard Error

**Table 3.** Longitudinal linear mixed-effects model of subcortical regional Volumes by estimated years to symptom onset interaction with mutation status.

| Subcortical region |  | Fixed-effects coefficient | SE | df | t-value | p-value |
| --- | --- | --- | --- | --- | --- | --- |
| Amygdala | Right | -6.64 | 2.34 | 346.00 | -2.83 | 0.005 |
|  | Left | -9.18 | 2.09 | 346.00 | -4.40 | **<0.001\*** |
| Thalamus | Right | -29.79 | 5.03 | 346.00 | -5.92 | **<0.001\*** |
|  | Left | -27.60 | 7.12 | 346.33 | -3.88 | **<0.001\*** |
| Putamen | Right | -24.45 | 5.59 | 346.00 | -4.37 | **<0.001\*** |
|  | Left | -22.17 | 6.12 | 346.00 | -3.62 | **<0.001\*** |
| Pallidum | Right | -0.67 | 1.46 | 346.00 | -0.46 | 0.645 |
|  | Left | -0.67 | 1.46 | 346.00 | 0.20 | 0.844 |
| Nucleus Accumbens | Right | -3.31 | 0.93 | 346.00 | -3.55 | **<0.001\*** |
|  | Left | -4.56 | 1.16 | 346.00 | -3.93 | **<0.001\*** |
| Hippocampus | Right | -26.35 | 4.49 | 346.00 | -5.86 | **<0.001\*** |
|  | Left | -23.53 | 4.46 | 346.00 | -5.27 | **<0.001\*** |
| Caudate | Right | -3.18 | 3.69 | 346.00 | -0.86 | 0.390 |
|  | Left | -2.33 | 3.47 | 346.00 | -0.67 | 0.502 |

Mutation status are divided by mutation carriers and non-carriers of autosomal dominant
Alzheimer's disease \*p < 0.05/14; SE = Standard Error

**Table 4.**
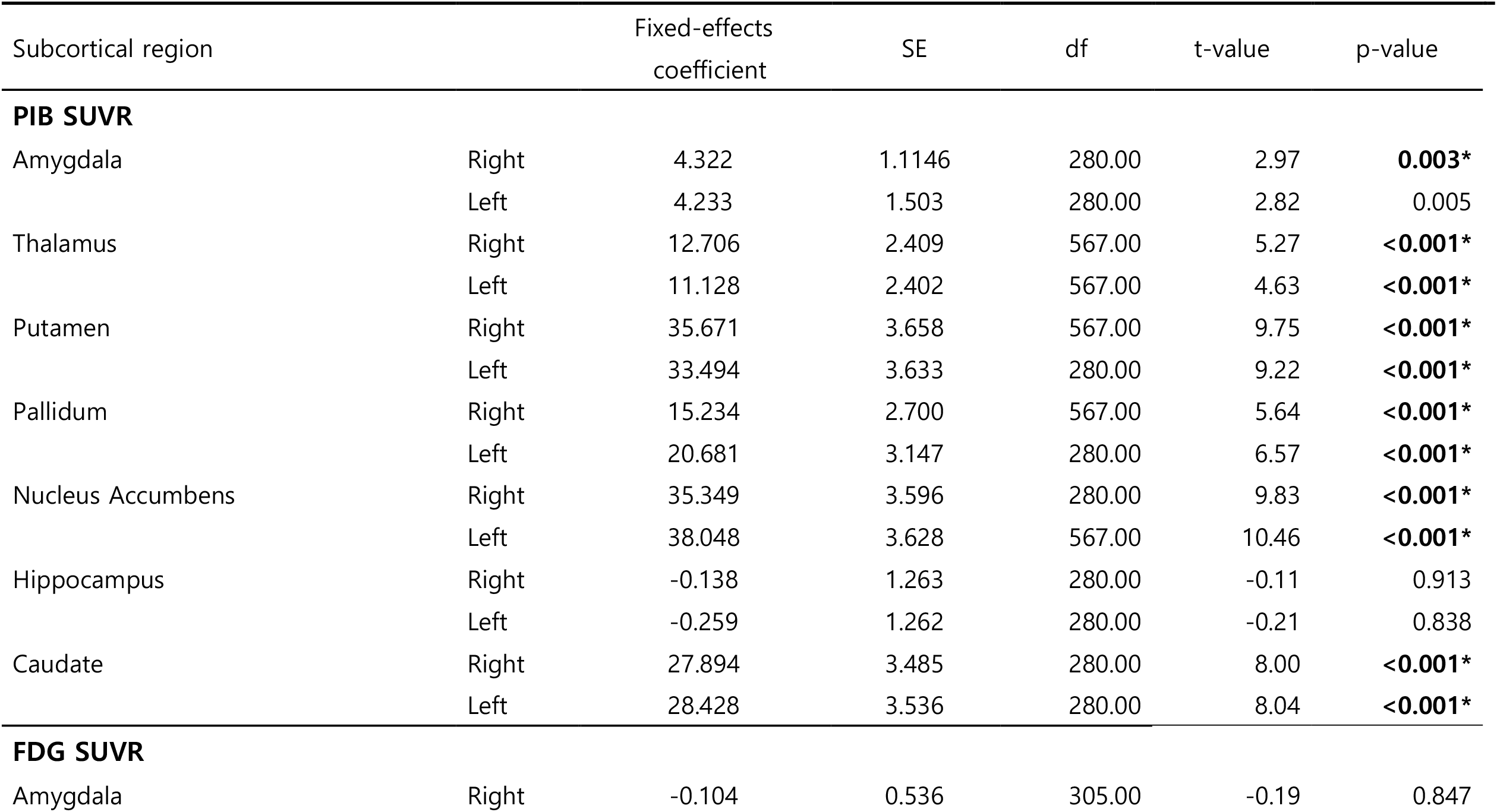
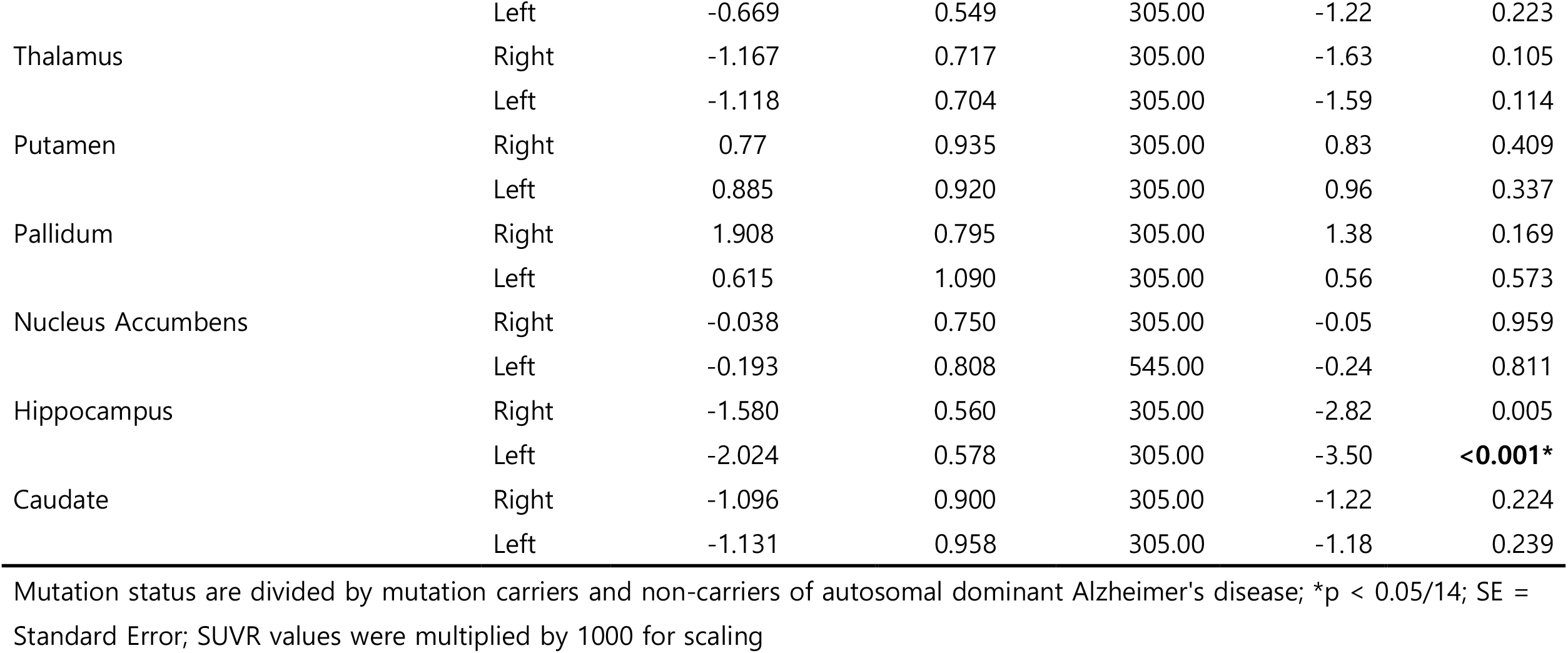
Longitudinal linear mixed-effects model of subcortical regional PIB/FDG SUVR by estimated years to symptom onset interaction with mutation status.

**Figure 1.**
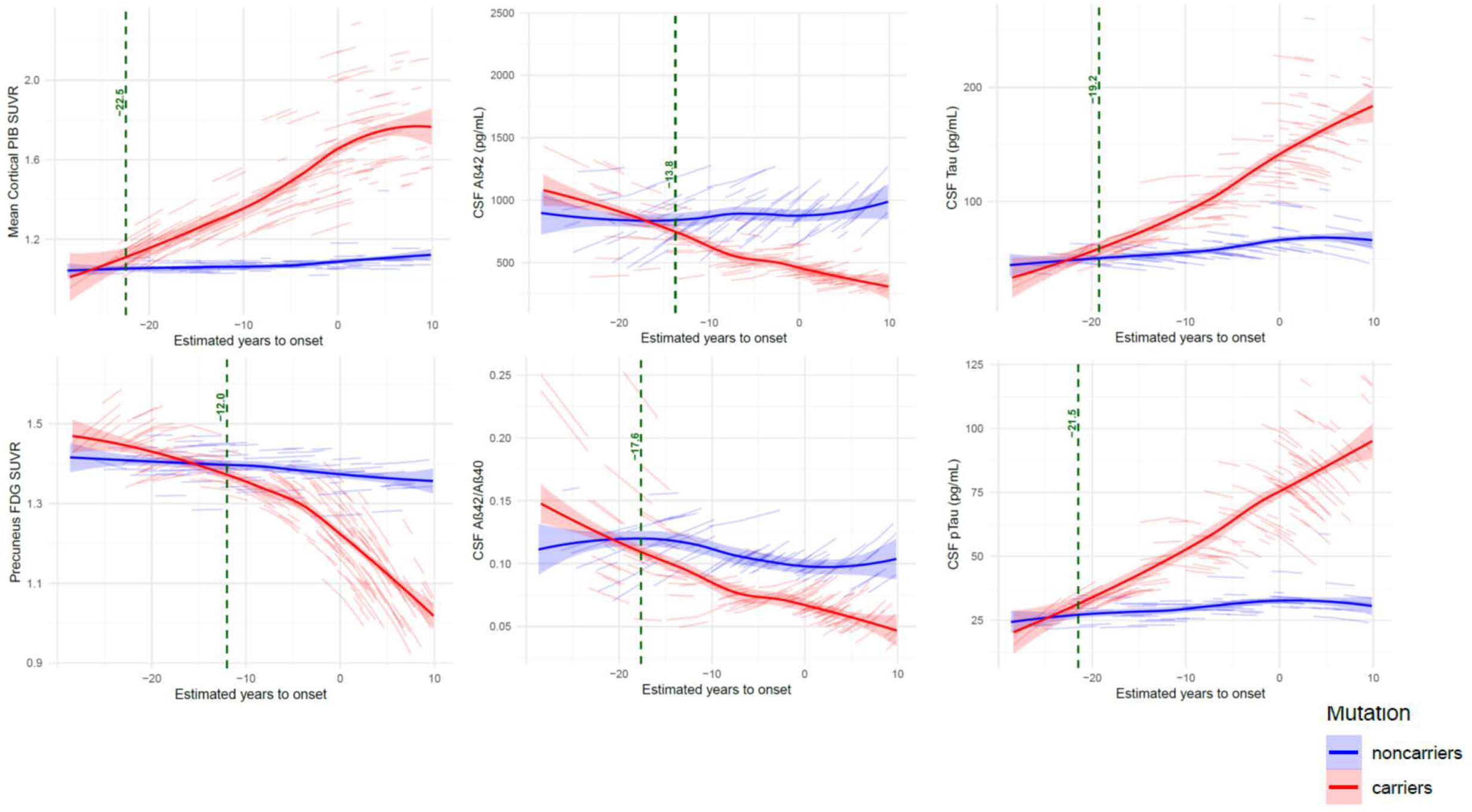
Longitudinal linear mixed-effects model of the reference AD biomarkers by estimated years to symptom onset interaction with mutation status. Dotted green line and text are starting estimated years to onset point with non-overlap zone of 99% confidence interval of predicted lines for mutation carriers and noncarriers. These dotted green lines are displayed only when a significant interaction between estimated years to symptom onset and mutation status is observed.

**Figure 2.**
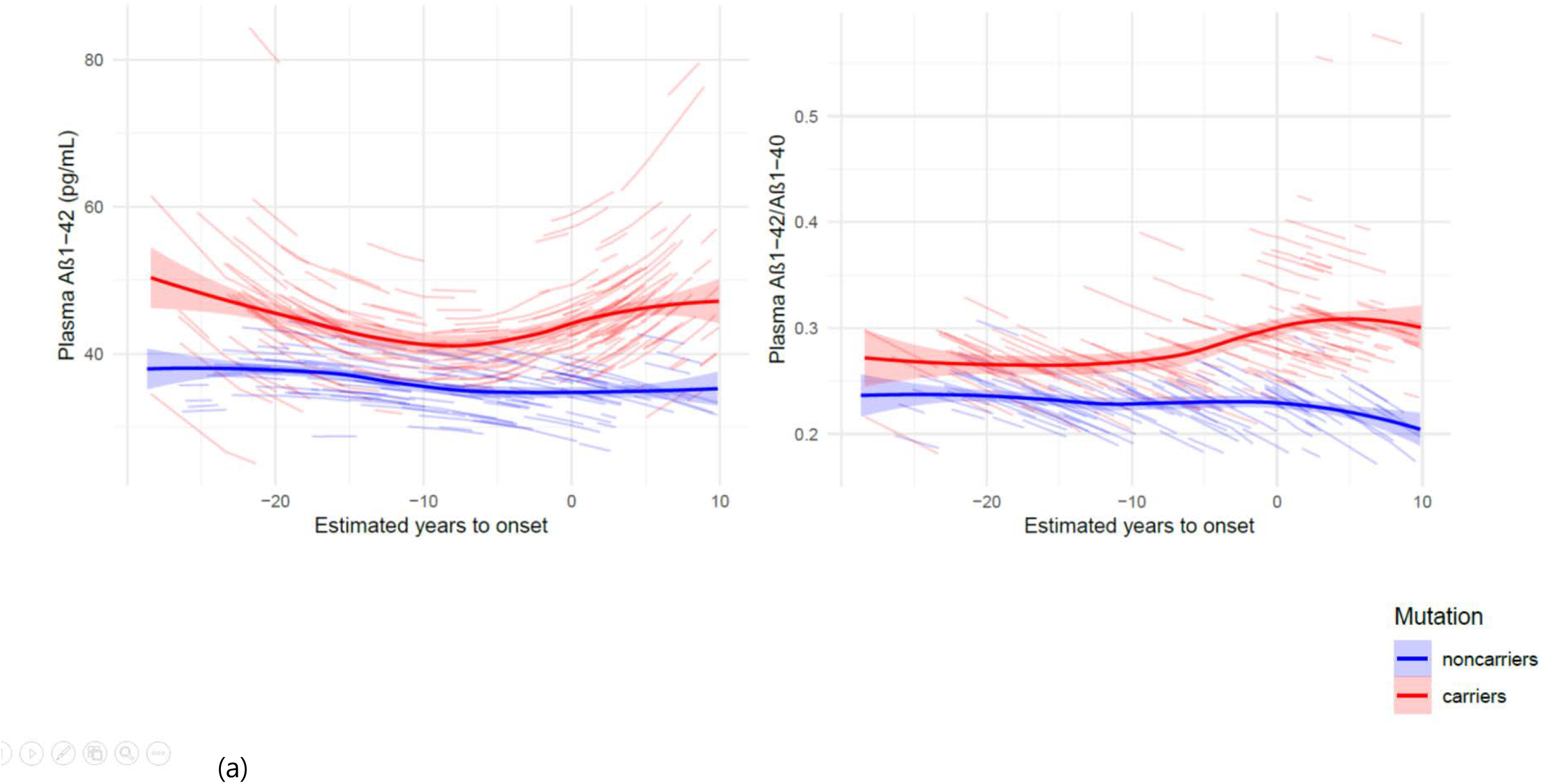
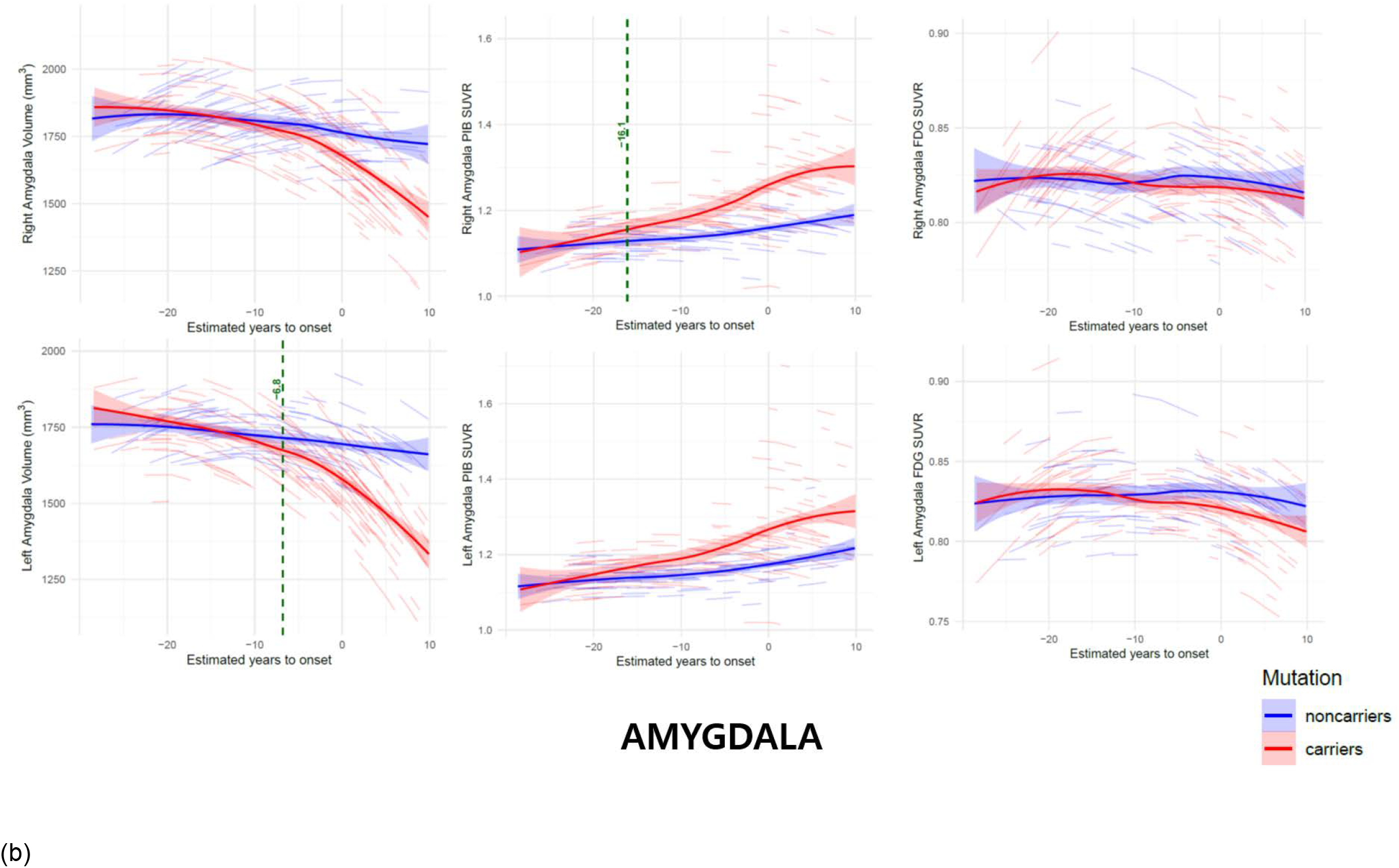
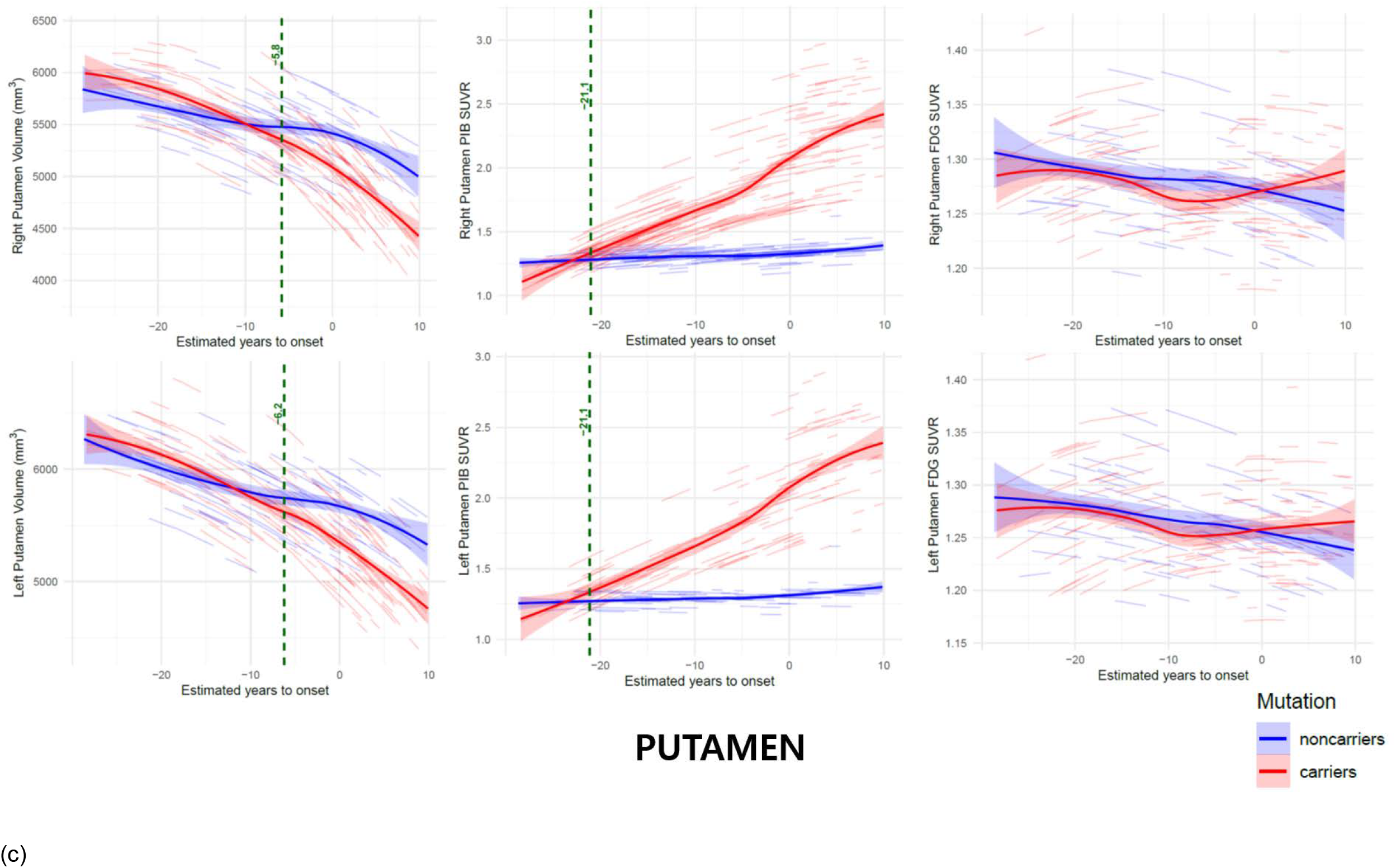
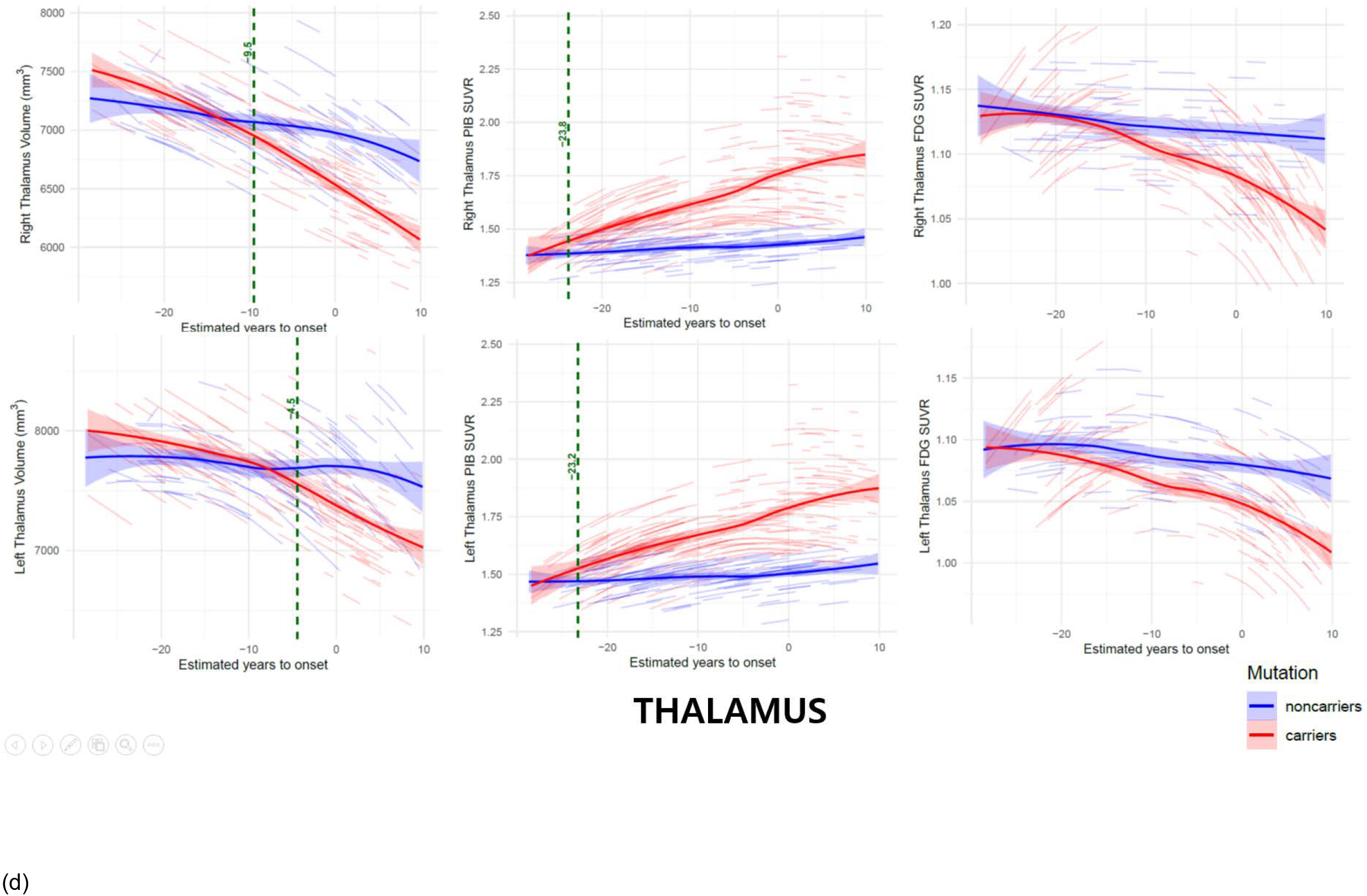
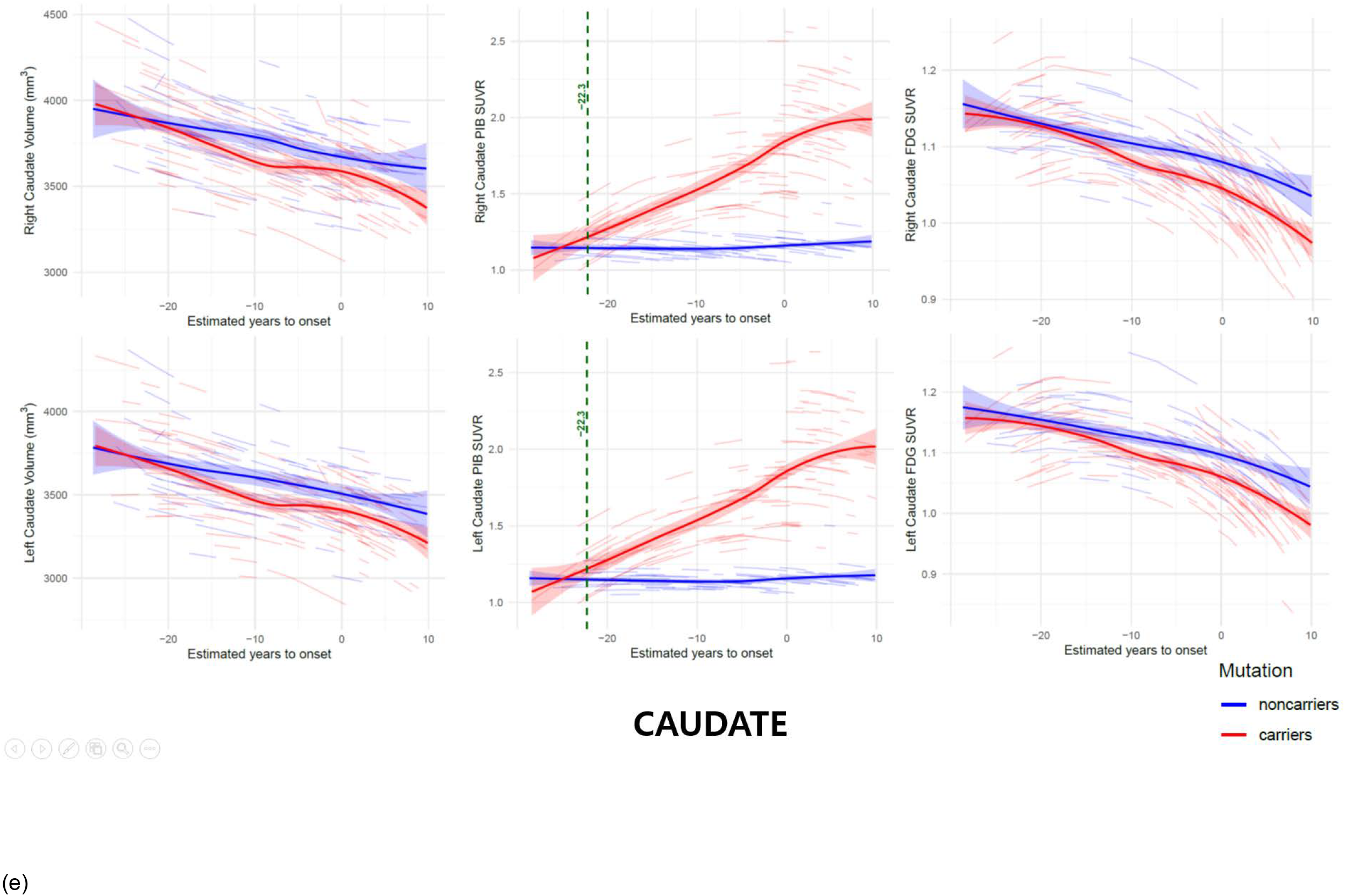
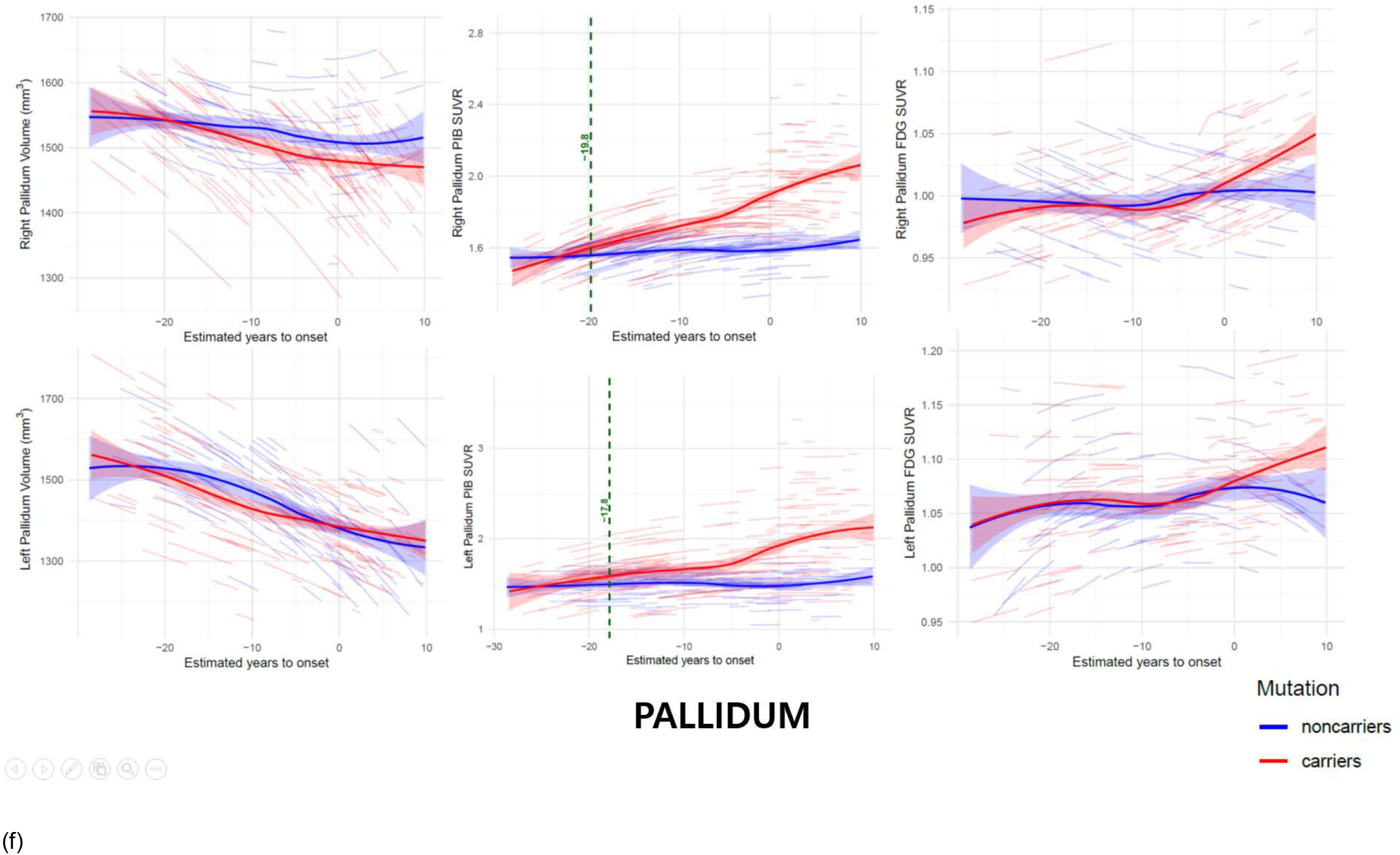
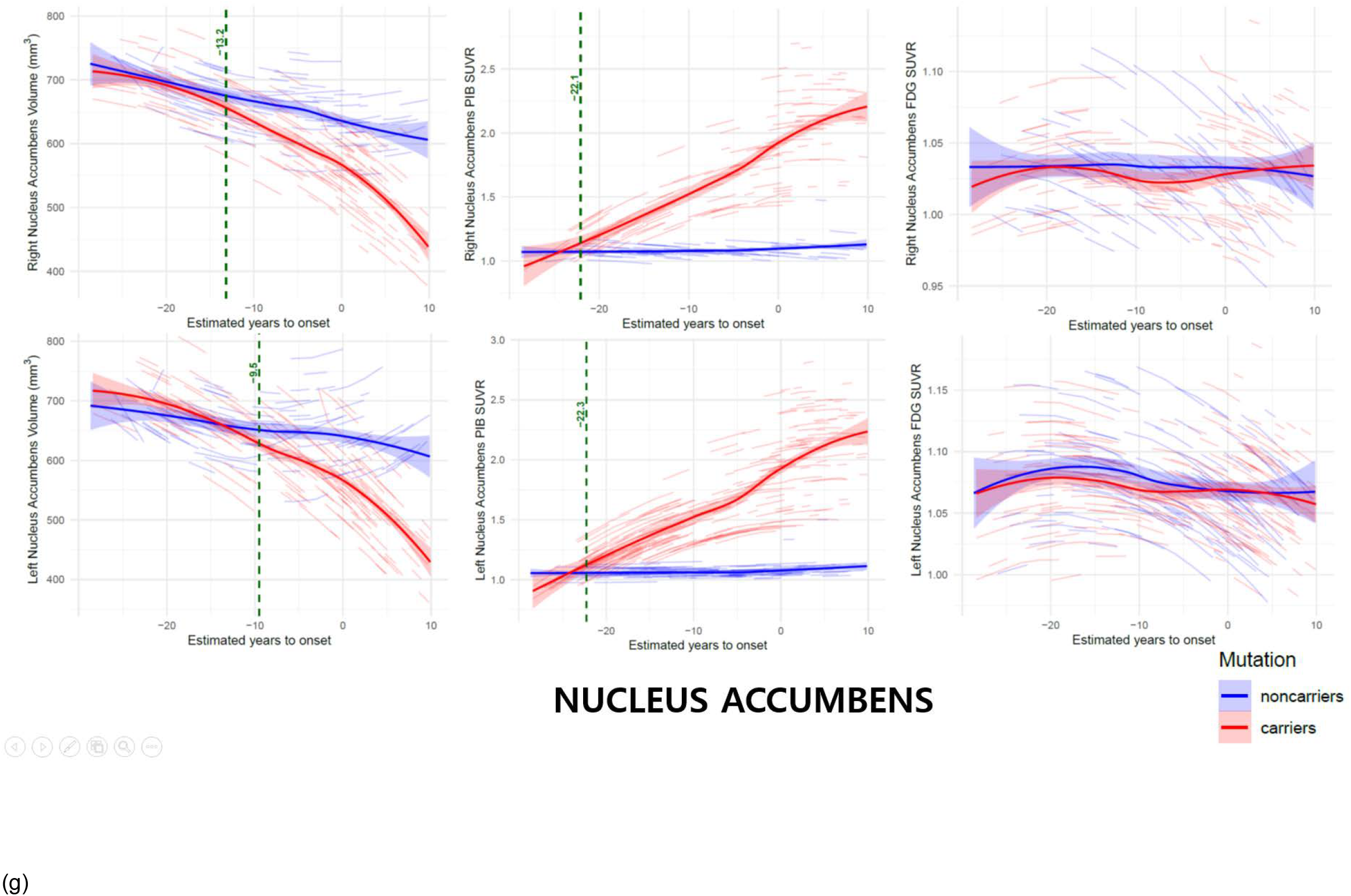
Longitudinal linear mixed-effects model of the subcortical regions by estimated years to symptom onset interaction with mutation status. (a) Amygdala (b) Putamen (c) Thalamus (d) Caudate (e) Pallidum (f) Nucleus Accumbens (g) Hippocampus. Left columns are volume, middle columns are PIB SUVRs, and right columns are FDG SUVRs. The dotted green line and accompanying text indicate the estimated time point at which the 99% confidence intervals of the predicted trajectories for mutation carriers and non-carriers no longer overlap. These dotted green lines are displayed only when a significant interaction between estimated years to symptom onset and mutation status is observed.

### Longitudinal Associations Between Defined Subcortical Volumes and AD Biomarkers in Mutation Carriers

The defined subcortical volumes included ten regions of interest (ROIs) — the bilateral amygdala, thalamus, putamen, nucleus accumbens, and hippocampus — which showed significant results in the first-step analyses. The right amygdala was also included due to a significant trend (p < 0.005).

Longitudinal association analyses were conducted exclusively in mutation carriers. Significant longitudinal associations between each subcortical volume and the region-matched PIB SUVR were observed in the bilateral amygdala, right thalamus, bilateral putamen, and bilateral nucleus accumbens (Supplementary Data Table 1, Supplementary Data Figure 1a). Regarding associations with region-matched FDG SUVR, significant associations were found in the left amygdala, left thalamus, left nucleus accumbens, and bilateral hippocampus (Supplementary Data Table 1, Supplementary Data Figure 1b). In addition, significantly negative longitudinal associations were identified between mean cortical PIB SUVR and subcortical volumes, except for the left thalamus (Supplementary Data Table 2), whereas significantly positive longitudinal associations were observed between the ten subcortical volumes and precuneus FDG SUVR (Supplementary Data Table 3).

For the analyses examining longitudinal associations between subcortical volumes and fluid AD biomarkers: Significant associations with CSF Aβ42 were found in the left amygdala, right thalamus, bilateral putamen, bilateral nucleus accumbens, and bilateral hippocampus. Significant associations with CSF Aβ42/Aβ40 ratio were observed in the right thalamus, bilateral putamen, bilateral nucleus accumbens, and left hippocampus. Significant associations with CSF Tau were present across all ten defined subcortical ROIs. Significant associations with CSF pTau were found in the left amygdala, right thalamus, bilateral putamen, bilateral nucleus accumbens, and bilateral hippocampus. For plasma biomarkers, significant associations with plasma Aβ1-42 were identified in the right amygdala and right hippocampus, and associations with plasma Aβ1-42/Aβ1-40 ratio were found in the right amygdala, right thalamus, and bilateral hippocampus (Supplementary Data Table 4).

## DISCUSSION

In the present study, we applied the linear-mixed effects model to reference AD biomarkers, to compare longitudinal trajectories between mutation carriers and non-carriers. Among the biomarkers, mean cortical Aβ deposition was the earliest biomarker to show divergence, with carriers exhibiting significantly greater accumulation beginning 22.5 years before the estimated age of symptom onset. This was followed by divergence in CSF pTau (21.5 years before), CSF Tau (19.2 years), CSF Aβ42/Aβ40 ratios (17.6 years), and CSF Aβ42 (13.8 years). In contrast, precuneus glucose metabolism diverged closer to symptom onset, at 12.0 years prior.

Compared to a previous DIAN longitudinal study,^8^ the timepoint of Aβ divergence was similar (22.2 vs. 22.5 years before). However, our finding on glucose metabolism divergence (12.0 years) occurred substantially closer to symptom onset than reported previously (18.8 years). The divergence timepoints for CSF Tau and CSF Aβ42 (19.2 and 13.8 years before onset, respectively) are consistent with prior cross-sectional analyses from the DIAN study (15 and 10 years, respectively),^23^ as well as with longitudinal findings from the same cohort,^24^ which reported divergence of CSF Tau at 17 years and CSF pTau at 19 years before onset. These prior findings align well with the present results (CSF Tau: 19.2 years; CSF pTau: 21.5 years before onset). Furthermore, this study adds novel longitudinal evidence by identifying an earlier divergence in the CSF Aβ42/Aβ40 ratio at 17.6 years before onset, reinforcing the temporally ordered cascade of pathological changes.

For subcortical regional analyses, we first observed that mutation carriers exhibited significantly greater longitudinal atrophy in the left amygdala, bilateral thalamus, putamen, nucleus accumbens, and hippocampus compared to non-carriers. Furthermore, carriers showed significantly higher longitudinal Aβ accumulation in most subcortical regions, except for the left amygdala and bilateral hippocampus. Regions with significant accumulation included the right amygdala, bilateral thalamus, putamen, pallidum, nucleus accumbens, and caudate and greater hypometabolic in left hippocampus relative to non-carriers.

Among subcortical imaging biomarkers, the earliest significant difference in volume was observed in the right nucleus accumbens, occurring 13.2 years before estimated onset, followed by the left nucleus accumbens (9.5 years) and right thalamus (9.5 years). For Aβ accumulation, the right thalamus showed the earliest divergence (23.8 years before onset), followed by the left thalamus (23.2 years), and then the left nucleus accumbens and bilateral caudate (22.3 years), all of which are comparable to the timing of mean cortical Aβ deposition (22.5 years before onset). In contrast, only the left hippocampus showed a significant decline in glucose metabolism between mutation carriers and non-carriers, with divergence occurring 8.1 years before the expected symptom onset. These findings suggest a distinct temporal sequence in subcortical regions, where β-amyloidosis precedes volumetric atrophy. This pattern differs from the previously proposed temporal cascade observed in cortical regions such as the precuneus, where Aβ accumulation is followed by metabolic decline and then cortical thinning. This interpretation is supported by our region-specific second-step analyses.

In our second-step analysis, subcortical regions exhibiting significant longitudinal atrophic changes were further examined for their associations with regional Aβ accumulation and glucose metabolism. Among mutation carriers, all of these regions-except the left thalamus and bilateral hippocampus-demonstrated significant associations with region-matched Aβ accumulation. In addition, region-matched glucose metabolism was significantly associated with volume changes in the left amygdala, left thalamus, left nucleus accumbens, and bilateral hippocampus. These subcortical volumes also showed significant associations with reference AD biomarkers, including mean cortical Aβ deposition, precuneus metabolism, and CSF Tau levels in mutation carriers. When considered alongside CSF biomarker results, longitudinal atrophy of the right thalamus, bilateral putamen, and bilateral nucleus accumbens was associated with either region-specific or mean cortical Aβ accumulation, as well as with CSF Aβ42, Aβ42/Aβ40 ratio, total Tau, and pTau. These findings suggest that longitudinal atrophy in these five subcortical ROIs reflects ongoing amyloid and tau pathophysiology in the context of dominantly inherited Alzheimer’s disease. Although tau PET imaging was not included in this study, such data could further elucidate region-specific tau pathology in these subcortical structures.

Regarding plasma biomarkers, although they had no significant interaction between mutation status and estimated years to symptom onset, in mutation carriers plasma Aβ42 levels were associated with volumes of the right amygdala and right hippocampus. Additionally, the plasma Aβ42/Aβ40 ratio was associated with the right amygdala, right thalamus, and bilateral hippocampus. These findings suggest that subcortical atrophy in certain regions may be related to plasma Aβ biomarker in mutation carriers.

The current work had a priori laterality predictions in subcortical regions, although not in the previous DIAN study.^8^ The laterality of longitudinal atrophy is prominent in the right thalamus. Although hemispheric differences of subcortical atrophy including thalamus have been noted in sporadic Alzheimer’s disease,^25-27^ our results provide additional laterality information of the longitudinal trajectories and divergence timepoints between mutation groups for subcortical AD biomarkers.

To our knowledge, the present study represents the most comprehensive and longest longitudinal analysis of biomarker data in autosomal dominant AD to date, with a specific focus on subcortical brain regions. However, the temporal sequence and relationships among biomarkers should be interpreted with caution, as individual-level data may not capture the full course of disease progression, and some individuals may deviate from the overall population trends.

Our findings suggest a distinct biomarker trajectory in subcortical regions, differing from the previously established cortical sequence of β-amyloidosis, followed by glucose hypometabolism, and subsequent structural atrophy. Specifically, β-amyloidosis appears first, and followed by subcortical volume atrophy. Moreover, subcortical volume loss is significantly associated with AD pathology. Given that MRI is more cost-effective and less time-consuming than PET in clinical settings, measuring subcortical volume atrophy through MRI may contribute to the early detection of Alzheimer’s disease.

## Supporting information

https://dian.wustl.edu/our-research/for-investigators/dian-observational-study-investigator-resources/data-request-terms-and-instructions/

## Data Availability

All datasets described within the current resource manuscript are freely available upon the completion of a Dominantly Inherited Alzheimer Network Data Request, (https://dian.wustl.edu/our-research/for-investigators/dian-observational-study-investigator-resources/data-request-terms-and-instructions/). Imaging data is available as extracted averages from FreeSurfer derived regions of interest (.xlsx), or identity-stripped source files (DICOM). Specifically, requesting the following data points from DIAN data release 16: MRI: Intracranial volume, subcortical (amygdala, thalamus, putamen, nucleus accumbens, hippocampus, caudate) volume; PET partial volume corrected SUVRs for: FDG-PET, PiB-PET summary regions; Demographics: Age, Sex, Education; Clinical/Cognition: CDR, MMSE, WAIS, delayed logical memory, Animal naming task, Boston naming task; Genetics: family ID, mutation carrying status, ADAD mutation type, age of expected symptom onset.

## Supplementary Materials

The Supplement is available with this article.

## Availability of Data and Material

The data used in this analysis are available on request to DIAN, provided data request applications are approved by the studies’ committees. Data for DIAN can be requested at https://dian.wustl.edu/forinvestigators/.

## Author Contributions

IH.C. designed the study, performed the analyses and wrote the initial draft of the manuscript. H.P. performed the statistical analyses. DIAN Consortium collected samples and data. All authors helped to interpret the results and reviewed drafts of the manuscript

## Acknowledgments

This manuscript has been reviewed by DIAN Study investigators for scientific content and consistency of data interpretation with previous DIAN Study publications. We acknowledge the altruism of the participants and their families and contributions of the DIAN research and support staff at each of the participating sites for their contributions to this study.

---

**Supplementary Figure 1.**
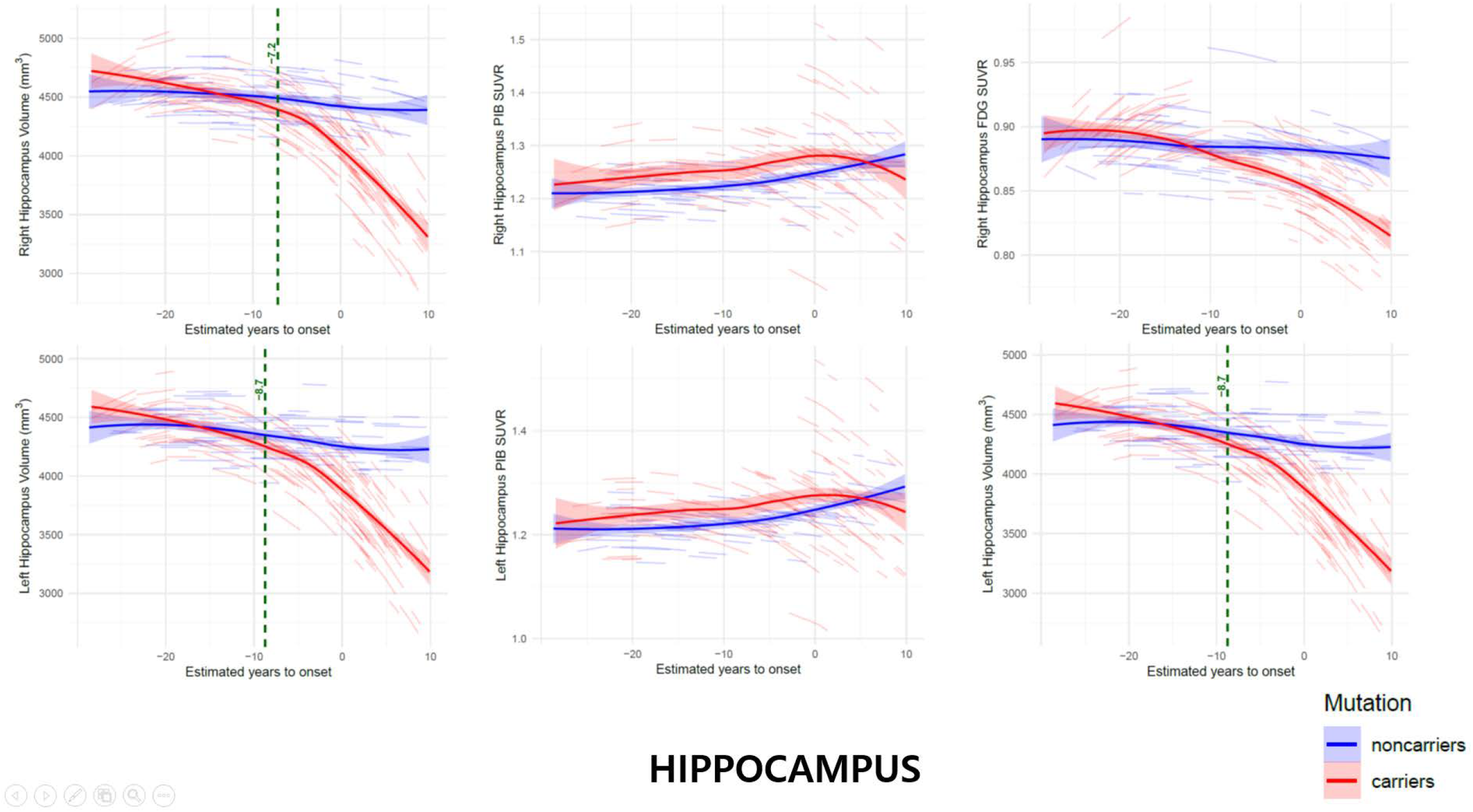
Significant longitudinal associations in the mutation carriers (a) between region-matched subcortical volumes and PIB SUVRs, and (b) between region-matched subcortical volumes and FDG SUVRs

