## Supplementary material for "Longitudinal subcortical volume changes and their correlations with multiple PET and fluid biomarkers in dominantly inherited Alzheimer’s disease": https://dian.wustl.edu/our-research/for-investigators/dian-observational-study-investigator-resources/data-request-terms-and-instructions/: DIAN-D2404_Supplementary Table_Figure.pdf

**Supplementary Table 1. Longitudinal associations between volume and PIB/FDG uptake in 10 subcortical regions of interest in mutation carriers.**

| Region | | Fixed-effects coefficient | SE | df | t-value | p-value | Correlation of Fixed Effects | $R^2_c$ |
|---|---|---|---|---|---|---|---|---|
| **PIB SUVR ~ Volume** | | | | | | | | |
| Amygdala | Right | -0.063 | 0.020 | 575.1 | -3.218 | **<0.001** | -0.955 | 0.89 |
| | Left | -0.079 | 0.022 | 601.7 | -3.572 | **<0.001** | -0.961 | 0.87 |
| Thalamus | Right | -0.044 | 0.015 | 548.7 | -2.926 | **<0.001** | -0.983 | 0.75 |
| | Left | -0.006 | 0.011 | 521.3 | -0.552 | 0.581 | -0.975 | 0.89 |
| Putamen | Right | -0.181 | 0.022 | 546.2 | -8.341 | **<0.001** | -0.963 | 0.94 |
| | Left | -0.117 | 0.018 | 490.6 | -6.589 | **<0.001** | -0.952 | 0.95 |
| Nucleus Accumbens | Right | -1.043 | 0.119 | 527.7 | -8.745 | **<0.001** | -0.919 | 0.94 |
| | Left | -0.416 | 0.098 | 459.7 | -4.249 | **<0.001** | -0.867 | 0.94 |
| Hippocampus | Right | 0.010 | 0.010 | 624.1 | 0.984 | 0.326 | -0.980 | 0.86 |
| | Left | -0.004 | 0.010 | 626.8 | -0.405 | 0.686 | -0.979 | 0.85 |
| **FDG SUVR ~ Volume** | | | | | | | | |
| Amygdala | Right | 0.010 | 0.008 | 606.2 | 1.227 | 0.220 | -0.978 | 0.61 |
| | Left | 0.030 | 0.009 | 599.4 | 3.393 | **0.001** | -0.98 | 0.58 |
| Thalamus | Right | 0.002 | 0.005 | 651.4 | 0.315 | 0.753 | -0.992 | 0.72 |
| | Left | -0.019 | 0.003 | 649.0 | -5.843 | **<0.001** | -0.986 | 0.74 |
| Putamen | Right | 0.004 | 0.006 | 631.6 | 0.664 | 0.507 | -0.984 | 0.75 |
| | Left | 0.002 | 0.005 | 653.5 | 0.446 | 0.656 | -0.982 | 0.75 |
| Nucleus Accumbens | Right | -0.018 | 0.026 | 625.0 | -0.684 | 0.494 | -0.967 | 0.68 |

| | | | | | | | | |
|---|---|---|---|---|---|---|---|---|
| | Left | 0.122 | 0.023 | 654.5 | 5.331 | **<0.001** | -0.955 | 0.66 |
| Hippocampus | Right | 0.043 | 0.004 | 489.0 | 10.25 | **<0.001** | -0.985 | 0.71 |
| | Left | 0.041 | 0.004 | 482.7 | 9.211 | **<0.001** | -0.984 | 0.72 |

Associations that were significant after correction for multiple comparisons using Bonferroni are indicated in bold; SE = standard error; $R^2c$ = conditional coefficient of determination; Each volume was scaled by dividing by 1000

**Supplementary Table 2. Longitudinal associations between the 10 subcortical regions of interest volumes and Mean Cortical PIB SUVR in mutation carriers**

| Region | | Fixed-effects coefficient | SE | df | t-value | p-value | Correlation of Fixed Effects | $R^2_c$ |
|---|---|---|---|---|---|---|---|---|
| Amygdala | Right | -0.123 | 0.033 | 466.2 | -3.748 | **<0.001** | -0.926 | 0.946 |
| | Left | -0.124 | 0.035 | 463.3 | -3.544 | **<0.001** | -0.928 | 0.945 |
| Thalamus | Right | -0.083 | 0.014 | 458.0 | -5.856 | **<0.001** | -0.973 | 0.946 |
| | Left | -0.015 | 0.010 | 429.0 | -1.520 | **0.129** | -0.953 | 0.949 |
| Putamen | Right | -0.092 | 0.015 | 537.5 | -6.16 | **<0.001** | -0.962 | 0.946 |
| | Left | -0.073 | 0.013 | 485.9 | -5.784 | **<0.001** | -0.951 | 0.948 |
| Nucleus Accumbens | Right | -0.471 | 0.081 | 496.1 | -5.804 | **<0.001** | -0.906 | 0.948 |
| | Left | -0.305 | 0.061 | 440.8 | -5.025 | **<0.001** | -0.845 | 0.948 |
| Hippocampus | Right | -0.145 | 0.020 | 621.4 | -7.175 | **<0.001** | -0.968 | 0.943 |
| | Left | -0.159 | 0.020 | 611.0 | -8.096 | **<0.001** | -0.965 | 0.944 |

Associations that were significant after correction for multiple comparisons using Bonferroni are indicated in bold; SE = standard error; $R^2c$ = conditional coefficient of determination; Each volume was scaled by dividing by 1000

**Supplementary Table 3. Longitudinal associations between the 10 subcortical regions of interest volumes and Precuneus FDG SUVR in mutation carriers**

| Region | | Fixed-effects coefficient | SE | df | t-value | p-value | Correlation of Fixed Effects | $R^2_c$ |
|---|---|---|---|---|---|---|---|---|
| Amygdala | Right | 0.21 | 0.02 | 624.7 | 10.85 | **<0.001** | -0.964 | 0.863 |
| | Left | 0.26 | 0.02 | 628.9 | 12.65 | **<0.001** | -0.967 | 0.858 |
| Thalamus | Right | 0.08 | 0.01 | 549.6 | 8.437 | **<0.001** | -0.989 | 0.829 |
| | Left | 0.02 | 0.00 | 596.2 | 4.847 | **<0.001** | -0.983 | 0.822 |
| Putamen | Right | 0.12 | 0.01 | 660.0 | 15.22 | **<0.001** | -0.980 | 0.875 |
| | Left | 0.08 | 0.01 | 652.3 | 10.51 | **<0.001** | -0.978 | 0.850 |
| Nucleus Accumbens | Right | 0.59 | 0.05 | 657.1 | 12.35 | **<0.001** | -0.958 | 0.843 |
| | Left | 0.38 | 0.04 | 596.9 | 9.927 | **<0.001** | -0.929 | 0.851 |
| Hippocampus | Right | 0.18 | 0.01 | 627.7 | 18.77 | **<0.001** | -0.981 | 0.896 |
| | Left | 0.17 | 0.01 | 609.8 | 18.15 | **<0.001** | -0.980 | 0.888 |

Associations that were significant after correction for multiple comparisons using Bonferroni are indicated in bold; SE = standard error; $R^2c$ = conditional coefficient of determination; Each volume was scaled by dividing by 1000

**Supplementary Table 4. Longitudinal associations between volumes in 10 subcortical regions of interest and AD biofluid biomarkers in mutation carriers.**

| Region | | Fixed-effects coefficient | SE | df | t-value | p-value | Correlation of Fixed Effects | $R^2_c$ |
|---|---|---|---|---|---|---|---|---|
| CSF Aβ42 | | | | | | | | |
| Amygdala | Right | 64.29 | 48.46 | 392.36 | 1.327 | 0.185 | -0.97 | 0.88 |
| | Left | 127.88 | 47.09 | 361.15 | 2.716 | **0.001** | -0.96 | 0.88 |
| Thalamus | Right | 115.68 | 21.82 | 451.12 | 5.302 | **<0.001** | -0.99 | 0.87 |
| | Left | 20.87 | 14.69 | 358.75 | 1.420 | 0.156 | -0.98 | 0.88 |
| Putamen | Right | 123.17 | 21.23 | 471.77 | 5.801 | **<0.001** | -0.98 | 0.86 |
| | Left | 88.25 | 18.38 | 451.58 | 4.802 | **<0.001** | -0.98 | 0.86 |
| Nucleus Accumbens | Right | 455.39 | 121.16 | 473.33 | 3.758 | **<0.001** | -0.96 | 0.87 |
| | Left | 367.48 | 95.31 | 391.50 | 3.856 | **<0.001** | -0.93 | 0.87 |
| Hippocampus | Right | 88.89 | 26.25 | 471.31 | 3.387 | **0.001** | -0.98 | 0.87 |
| | Left | 76.42 | 25.21 | 467.67 | 3.031 | **0.003** | -0.98 | 0.87 |
| CSF Aβ42/40 | | | | | | | | |
| Amygdala | Right | 0.013 | 0.009 | 438.10 | 1.457 | 0.146 | -0.98 | 0.56 |
| | Left | 0.021 | 0.009 | 460.20 | 2.438 | 0.015 | -0.98 | 0.55 |
| Thalamus | Right | 0.016 | 0.004 | 397.45 | 4.411 | **<0.001** | -0.99 | 0.55 |
| | Left | 0.004 | 0.003 | 462.80 | 1.513 | 0.131 | -0.99 | 0.57 |
| Putamen | Right | 0.015 | 0.003 | 374.10 | 4.367 | **<0.001** | -0.99 | 0.55 |
| | Left | 0.014 | 0.003 | 404.80 | 4.482 | **<0.001** | -0.99 | 0.55 |
| Nucleus Accumbens | Right | 0.072 | 0.020 | 412.15 | 3.542 | **<0.001** | -0.98 | 0.56 |

| | | | | | | | | |
|---|---|---|---|---|---|---|---|---|
| | Left | 0.072 | 0.017 | 443.77 | 4.268 | **<0.001** | -0.97 | 0.55 |
| Hippocampus | Right | 0.011 | 0.004 | 348.70 | 2.798 | 0.005 | -0.99 | 0.56 |
| | Left | 0.013 | 0.004 | 369.90 | 3.164 | **0.002** | -0.99 | 0.55 |
| CSF Tau | | | | | | | | |
| Amygdala | Right | -47.71 | 12.00 | 471.94 | -3.976 | **<0.001** | -0.98 | 0.75 |
| | Left | -50.38 | 11.97 | 464.30 | -4.208 | **<0.001** | -0.98 | 0.73 |
| Thalamus | Right | -26.51 | 5.17 | 459.79 | -5.125 | **<0.001** | -0.99 | 0.73 |
| | Left | -11.35 | 3.73 | 457.17 | -3.041 | **0.002** | -0.99 | 0.76 |
| Putamen | Right | -31.11 | 4.91 | 431.67 | -6.334 | **<0.001** | -0.99 | 0.74 |
| | Left | -25.70 | 4.34 | 458.91 | -5.918 | **<0.001** | -0.99 | 0.74 |
| Nucleus Accumbens | Right | -177.33 | 28.65 | 462.12 | -6.189 | **<0.001** | -0.97 | 0.74 |
| | Left | -122.61 | 23.59 | 471.88 | -5.198 | **<0.001** | -0.96 | 0.73 |
| Hippocampus | Right | -43.46 | 5.67 | 387..505 | -7.659 | **<0.001** | -0.99 | 0.73 |
| | Left | -44.80 | 5.54 | 408.56 | -8.088 | **<0.001** | -0.98 | 0.72 |
| CSF pTau | | | | | | | | |
| Amygdala | Right | -6.49 | 5.78 | 470.22 | -1.122 | 0.262 | -0.98 | 0.75 |
| | Left | -19.56 | 5.77 | 462.57 | -3.389 | **0.001** | -0.98 | 0.73 |
| Thalamus | Right | -14.19 | 2.47 | 446.07 | -5.743 | **<0.001** | -0.99 | 0.72 |
| | Left | -2.92 | 1.80 | 457.40 | -1.624 | **0.105** | -0.99 | 0.75 |
| Putamen | Right | -13.53 | 2.35 | 422.31 | -5.766 | **<0.001** | -0.99 | 0.72 |
| | Left | -13.16 | 2.06 | 457.51 | -6.390 | **<0.001** | -0.99 | 0.74 |
| Nucleus Accumbens | Right | -79.29 | 13.68 | 457.82 | -5.795 | **<0.001** | -0.97 | 0.73 |
| | Left | -36.99 | 11.42 | 470.90 | -3.238 | **0.001** | -0.96 | 0.72 |

| | | | | | | | | |
|---|---|---|---|---|---|---|---|---|
| Hippocampus | Right | -17.06 | 2.76 | 384.54 | -6.194 | **<0.001** | -0.99 | 0.72 |
| | Left | -17.26 | 2.69 | 405.27 | -6.407 | **<0.001** | -0.98 | 0.70 |
| Plasma Aβ1-42 | | | | | | | | |
| Amygdala | Right | -6.58 | 2.29 | 586.92 | -2.877 | **0.004** | -0.98 | 0.74 |
| | Left | -5.42 | 2.27 | 584.28 | -2.387 | 0.017 | -0.97 | 0.74 |
| Thalamus | Right | -3.94 | 1.64 | 584.37 | -2.405 | 0.017 | -0.88 | 0.78 |
| | Left | -1.30 | 0.71 | 569.55 | -1.833 | 0.067 | -0.99 | 0.74 |
| Putamen | Right | -0.24 | 1.01 | 521.17 | -0.240 | 0.810 | -0.99 | 0.74 |
| | Left | 0.50 | 0.87 | 576.21 | 0.580 | 0.562 | -0.98 | 0.74 |
| Nucleus Accumbens | Right | -2.86 | 5.76 | 568.77 | -0.497 | 0.620 | -0.97 | 0.74 |
| | Left | 4.28 | 4.42 | 587.08 | 0.966 | 0.334 | -0.95 | 0.74 |
| Hippocampus | Right | -3.94 | 1.18 | 486.14 | -3.339 | **0.001** | -0.98 | 0.74 |
| | Left | -3.01 | 1.11 | 544.39 | -2.697 | 0.007 | -0.98 | 0.74 |
| Plasma Aβ1-42/Aβ1-40 | | | | | | | | |
| Amygdala | Right | -0.05 | 0.02 | 580.3 | -2.970 | **0.003** | -0.98 | 0.71 |
| | Left | -0.03 | 0.02 | 588.2 | -1.645 | 0.101 | -0.97 | 0.71 |
| Thalamus | Right | -0.03 | 0.01 | 569.7 | -4.156 | **<0.001** | -0.99 | 0.71 |
| | Left | -0.01 | 0.00 | 579.2 | -2.070 | 0.039 | -0.99 | 0.72 |
| Putamen | Right | -0.01 | 0.01 | 505.0 | -1.748 | 0.081 | -0.99 | 0.87 |
| | Left | -0.01 | 0.01 | 567.0 | -1.859 | 0.064 | -0.98 | 0.72 |
| Nucleus Accumbens | Right | -0.10 | 0.04 | 555.3 | -2.677 | 0.008 | -0.97 | 0.72 |
| | Left | -0.01 | 0.03 | 589.0 | -0.402 | 0.687 | -0.95 | 0.72 |
| Hippocampus | Right | -0.031 | 0.008 | 468.1 | -3.960 | **<0.001** | -0.99 | 0.71 |

| | | | | | | | |
|---|---|---|---|---|---|---|---|
| Left | -0.024 | 0.007 | 525.9 | -3.248 | **0.001** | -0.98 | 0.70 |

Associations that were significant after correction for multiple comparisons using Bonferroni are indicated in bold; SE = standard error; $R^2c$ = conditional coefficient of determination; Each volume was scaled by dividing by 1000

**Supplementary Figure 1** (a)

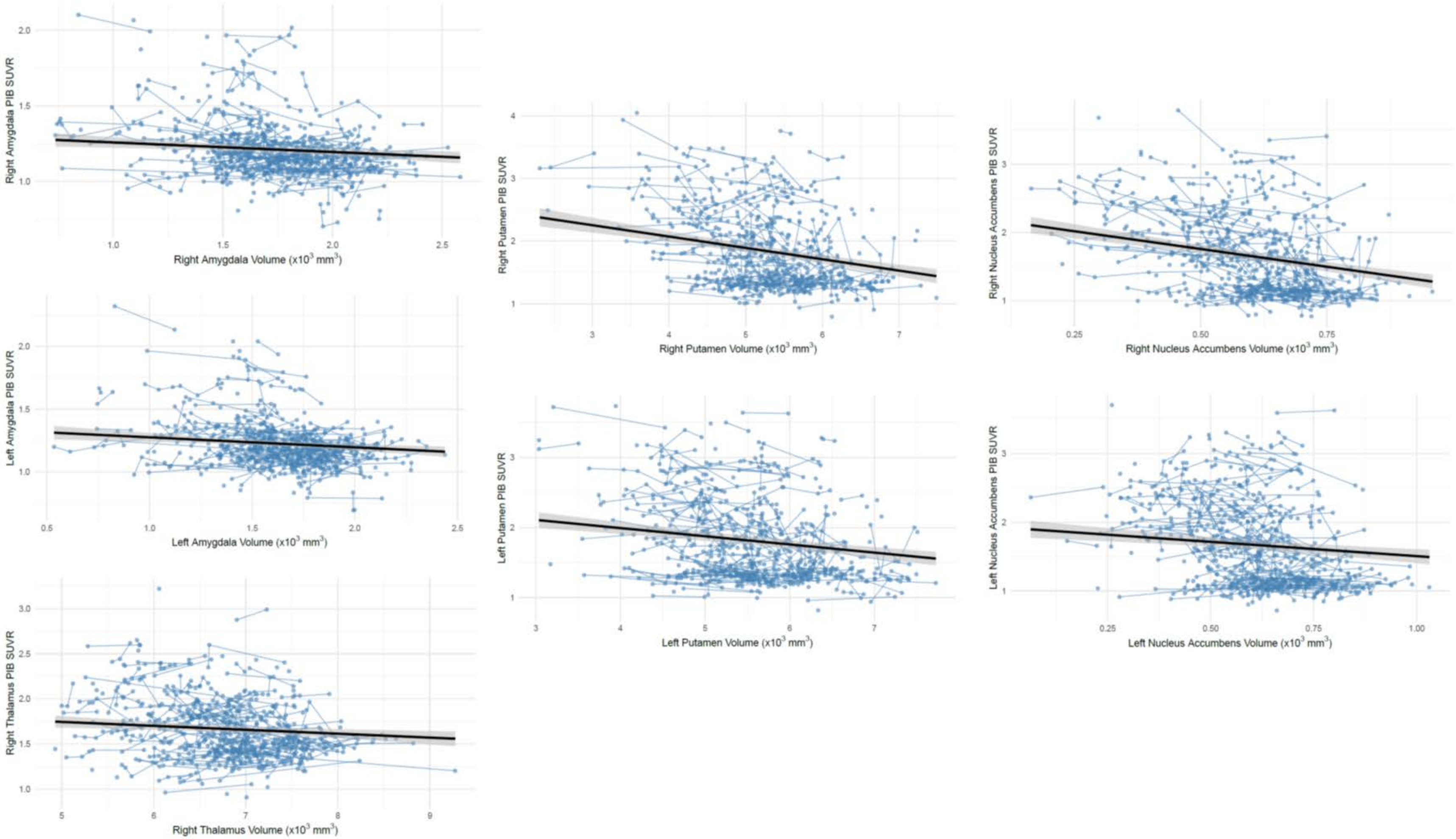

(b)

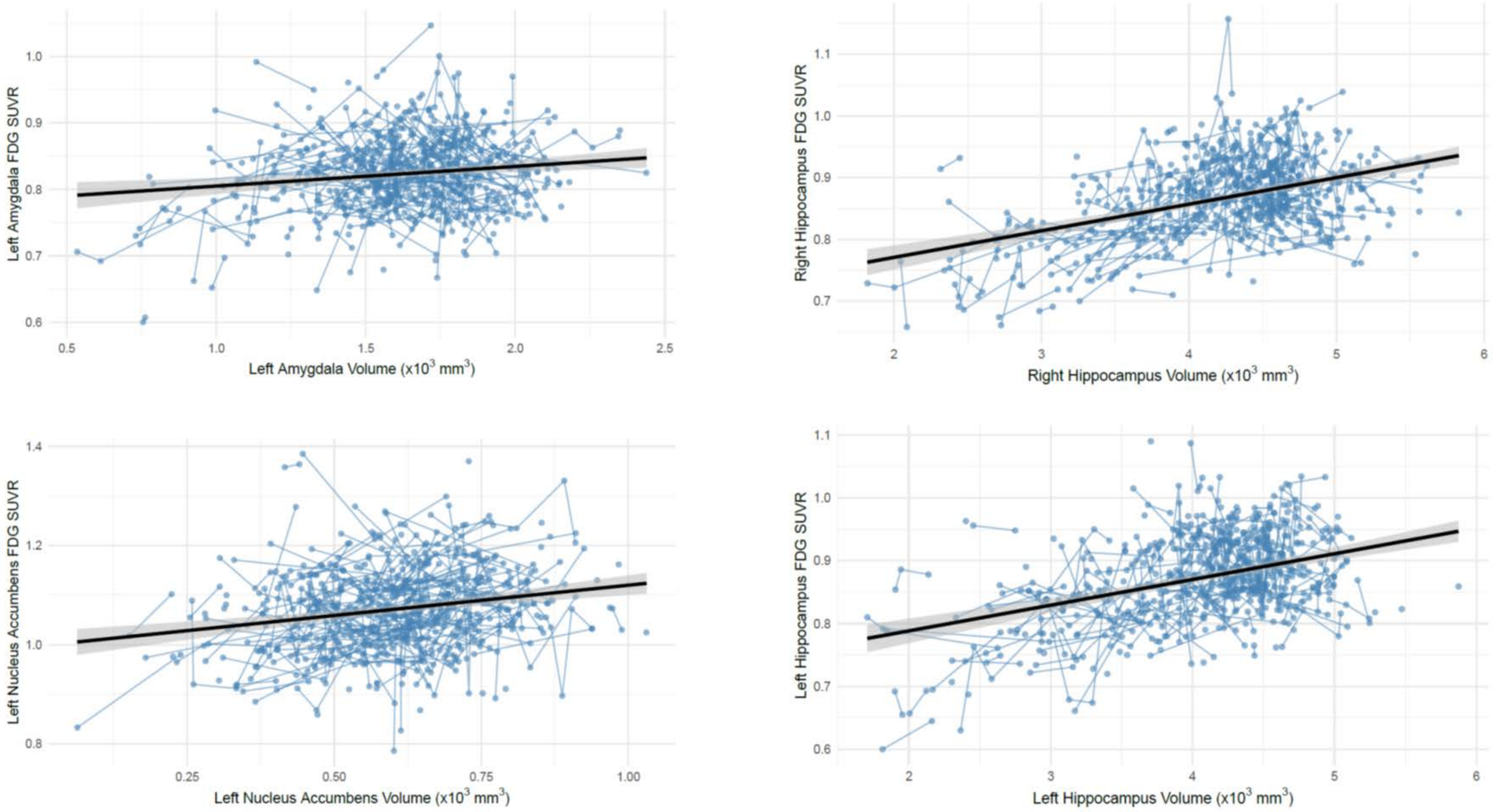
Left Amygdala FDG SUVR
Left Amygdala Volume (x10$^3$ mm$^3$)
Right Hippocampus FDG SUVR
Right Hippocampus Volume (x10$^3$ mm$^3$)
Left Nucleus Accumbens FDG SUVR
Left Nucleus Accumbens Volume (x10$^3$ mm$^3$)
Left Hippocampus FDG SUVR
Left Hippocampus Volume (x10$^3$ mm$^3$)
